# Humoral cross-reactivity towards SARS-CoV-2 in young children with acute respiratory infection with low-pathogenicity coronaviruses

**DOI:** 10.1101/2021.10.01.21264349

**Authors:** Nitin Dhochak, Tanvi Agrawal, Heena Shaman, Naseem Ahmed Khan, Prawin Kumar, Sushil K Kabra, Guruprasad R Medigeshi, Rakesh Lodha

## Abstract

SARS-CoV-2 infection in children frequently leads to only asymptomatic and mild infections. It has been suggested that frequent infections due to low-pathogenicity coronaviruses in children, imparts immunity against SARS-CoV-2 in this age group. From a prospective birth cohort study prior to the pandemic, we identified children (n=42) with proven low-pathogenicity coronavirus infections. Convalescent sera from these samples had antibodies against the respective seasonal CoVs as demonstrated by immunofluorescence assay. We tested these samples for neutralization of SARS-CoV-2 using virus microneutralization assay. Forty serum samples showed no significant neutralization of SARS-CoV-2, while 2 samples showed inconclusive results. These findings suggest that the antibodies generated in low-pathogenicity coronavirus infections offer no protection from SARS-CoV-2 infection in young children.

## Background

Young children frequently develop viral acute respiratory infections (ARI). Low-pathogenicity human coronaviruses (HCoVs) namely OC43, HKU1, NL63 and 229E have been reported in 6.5% of all ARI episodes in young children [1]. As compared to high morbidity and mortality in older population, SARS-CoV-2 infections in children have predominantly been mild or asymptomatic. Cross-immunity from early life common coronavirus infections towards SARS-CoV-2 has been postulated as a possible protective mechanism in COVID-19 in children. Cross-reactive antibodies to N-protein of SARS-CoV-2 have been reported in sera of up to 16.2% of individuals with common coronavirus infections in adults and children [2]. In another study, none of the 37 pre-pandemic sera with prior low-pathogenicity coronavirus infection showed cross-neutralization of SARS-CoV-2 [3]. Hence, we planned to study cross-neutralization of SARS-CoV-2 by convalescent serum of low-pathogenicity coronavirus infection in young children.

## Methods

From a prospective birth-cohort study evaluating the influence of viral infections in early life on pulmonary function at 3 years age, we identified children who had ARI due to coronavirus and had serum sample available within a year of documented infection; the methods and results of this cohort study in the pre-pandemic period are published [4]. Patients’ demographic details, species of coronavirus detected (OC43, HKU1, NL63 and 229E), and age at the time of ARI and serum collection were collated. The convalescent serum samples from children infected with HCoVs OC43, NL63 and 229E were evaluated for antibodies against respective human coronaviruses by immunofluorescence assay. One of the serum samples collected from healthy children from the same cohort who had no history of HCoV infection served as a negative control for the immunofluorescence assay. Anti-SARS-CoV-2 activity in preserved serum samples was assessed by virus microneutralization assay. The current study was conducted after obtaining clearance from Institute Ethics Committee with waiver of individual consent [IECPG 756/23.12.2020; RT-01/2020]

### Virus microneutralization assay

Vero E6 cells (NCCS, Pune, India) were maintained in DMEM high glucose medium (HiMedia) supplemented with 10% heat-inactivated FBS, 100U of penicillin and 100 µg of streptomycin in 5% CO_2_ incubator. SARS-CoV-2 THSTI-BAL-2010D was isolated and propagated as described elsewhere [5]. Vero E6 cells were seeded at 30,000 cells per well in a 96-well plate. Serum samples were heat-inactivated at 56°C for 30 min before use for this assay. 75 µl of heat-inactivated serum samples were two-fold diluted from 1:10 to 1:1280 using growth medium with 2% heat-inactivated FBS. To this, 75 µl of SARS-CoV-2 (final 1:16 dilution of 5 × 10^4^ PFU/ml stock) was added and kept for 1 h at 37°C in 5% CO_2_ incubator for virus neutralization. The virus-serum mixtures were then added on confluent monolayer of Vero E6 cells and incubated for 1 h at 37°C in 5% CO_2_ incubator for virus adsorption. After 1 h, viral inoculum was removed and overlaid with 2% carboxymethylcellulose in growth medium with 2% heat-inactivated FBS and incubated at 37°C in 5% CO_2_ incubator. At 24 h post infection, cells were fixed with 7.4% formaldehyde solution. Cells were washed 6 times with PBS and treated with 0.3% H_2_O_2_ for 20 minutes. Cells were then stained with anti-spike RBD antibody at 1:4000 dilution (Sino Biologicals; 40592-T62) for 1 h, followed by HRP-conjugated anti-rabbit antibody at 1:4000 dilution (Invitrogen; G-21234) for 1 h. After washing with PBS three times, 100 µl of TrueBlue substrate (Sera Care, 5510-0050) was added for 10 minutes. The microspots were quantified by AID iSPOT reader (ELR08IFL; AID GmbH, Strassberg, Germany) using AID EliSpot 8.0 software. 50% neutralization values were calculated with four-parameter logistic regression using GraphPad Prism 7.0e software. Pooled convalescent serum from COVID-19 patients was used as a positive control and pooled pre-pandemic sera or sera from COVID-19 antibody negative samples were used as a negative control with every assay run.

### Cell lines and virus for Immunofluorescences Assays

LLC MK2 cells were from European Collection of Authenticated Cell Cultures Salisbury, UK (ECACC - 85062804). Baby Hamster Kidney (BHK-21) cells were from American Type Culture Collection, Manassas, VA, USA (ATCC-CCL-10). Cells were grown at 37°C in Minimum Essential medium (MEM) (Gibco-11090-073) containing 10% heat-inactivated fetal bovine serum (FBS) (Gibco-16140-071), 100 U/mL of penicillin and 100 µg/mL of streptomycin (Gibco-10378-016). Human hepatoma cells (Huh-7) (Japanese Collection of Research Bioresources Cell Bank) were grown at 37° C in Dulbecco’s minimum essential medium (DMEM) (Lonza 12-707F) containing above additives and non-essential amino acids (Gibco-111400-50). Virus isolates were obtained through BEI Resources, NIAID, NIH: Human Coronavirus OC43, NR-52725; Human Coronavirus NL63, NR-470; Human Coronavirus 229E, NR-52726. Virus propagation and staining of infected cells using clinical samples has been described in Supplementary Material 1.

## Results

Of 310 children in the cohort, 42 children (21 male) were included in current study. Median (IQR) age at time of coronavirus ARI was 18 (14, 28) months. Time to blood sample collection after documented coronavirus ARI was 220 (174, 285) days (range: 21-347 days). Human coronaviruses OC43, HKU1, NL63 and 229E infections were seen in 11 (26%), 13 (31%), 10 (24%), and 8 (19%) children, respectively. Co-infection with other respiratory viruses was seen in 11 children (including another species of coronavirus in 3 children).

The respective serum samples were confirmed to be positive for the presence of antibodies against HCoVs OC43, NL63, and 229E by immunofluorescence assays (Supplementary Figures 1-3). The serum from a pediatric subject with no known history of HCoV infection did not show any staining while a positive sample and a positive control (commercial antibody) both showed specific staining (Supplementary Figure 4).

Forty of the HCoV convalescent serum samples showed no cross-reactive antibodies to SARS-CoV-2 as observed by the absence of neutralization in virus microneutralization assays while the in-house reference reagent (pooled convalescent serum from COVID-19 patients) showed neutralization of SARS-CoV-2 (Figure 1). The results were inconclusive in two samples.

**Figure 1.**
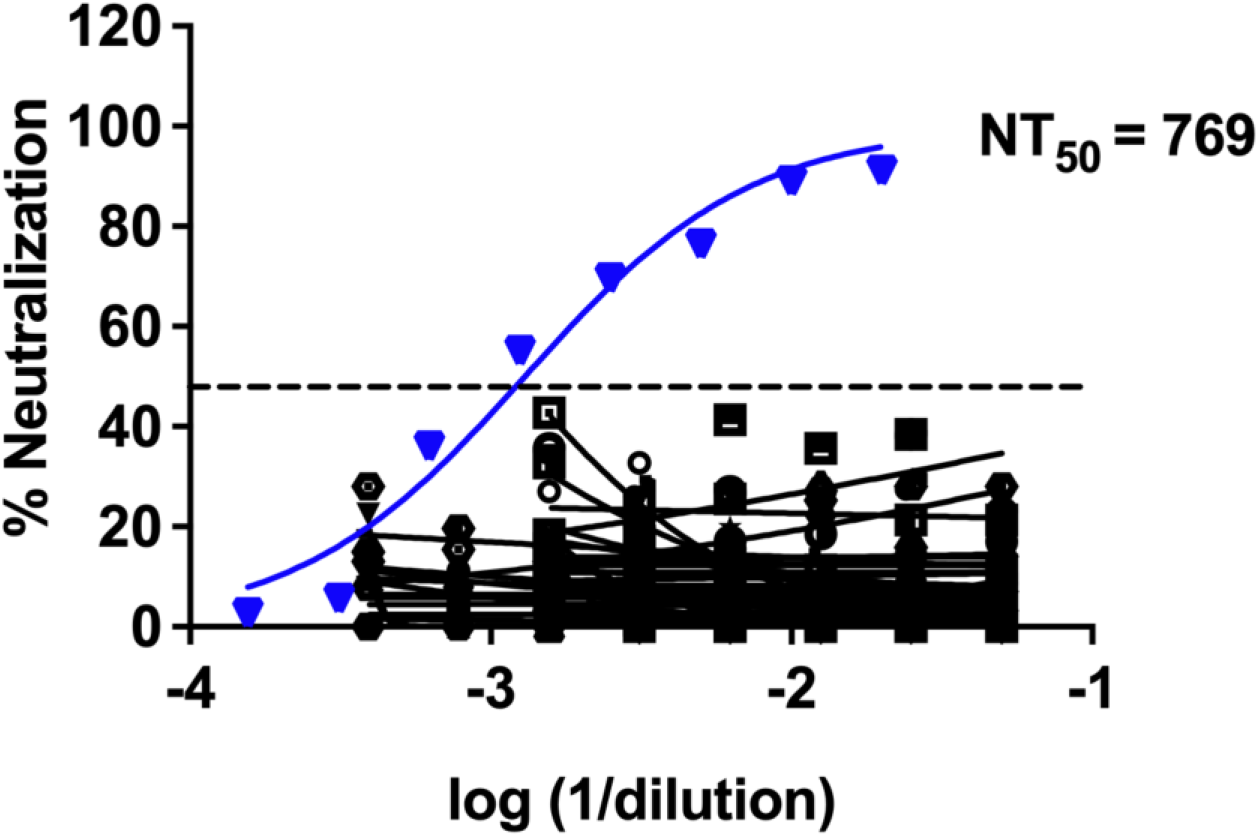
SARS-CoV-2 microneutralization assay: Serum samples from patients who had prior infection with seasonal coronaviruses was used to determine the titers of cross-neutralizing antibodies for SARS-CoV-2 virus by microneutralization assay. A reference reagent (pooled convalescent sera form COVID-19 patients) was calibrated against the WHO international reference reagent and was used as a positive control (**inverted triangle symbol**).

## Discussion

We observed that none of the young children aged less than 3 years infected with low-pathogenicity HCoVs demonstrated cross-neutralization against SARS-CoV-2 in convalescent sera, despite the presence of antibodies against individual coronaviruses.

In a previous study, following a low-pathogenicity coronavirus infection, 4.2%, 1%, and 16.2% samples from adults and children showed cross-reactive antibodies to S, RBD, and N proteins of SARS-CoV-2, respectively, as assessed by enzyme-linked immunosorbent assay; however, there was no cross neutralization with plaque reduction neutralization test (PRNT) [2]. We utilized wild-type virus microneutralization assay which is highly specific and has efficacy comparable to PRNT for neutralizing antibodies [6]. Even though our study points against the role of humoral cross-immunity against SARS-CoV-2 after infection with low-pathogenicity coronaviruses, cell-mediated cross-protection, which may contribute to lesser severity of SARS-CoV-2 infection in children, cannot be ruled out [7].

As we included a longer time period between ARI and serum sampling (up to 1 year of initial coronavirus infection), immunoglobulin levels in many children may have declined. While we could not perform assays for virus neutralization against the seasonal coronaviruses, we found that the respective serum samples used in this study were positive for antibodies against the three seasonal coronaviruses (OC43, NL63, 229E) by immunofluorescence. In addition, the presence of symptomatic ARI and PCR based detection of coronavirus ascertains coronavirus infection prior to serum collection. Also, in another study, neutralizing antibodies against human coronavirus OC43 were seen in 73% of convalescent sera, though none neutralized SARS-CoV-2 [3].

We conclude that infection with low-pathogenicity coronaviruses in children younger than 3 years does not impart humoral immunity against SARS-CoV-2. Cell mediated cross-immunity should be studied in this subgroup.

## Data Availability

The data may be shared on receiving the requests from investigators

## Footnote

### Funding source

The study was partly supported by grants from the Department of Biotechnology, Govt. of India, through IndCEPI Mission (BT/MB/CEPI/2016), Translational Research Program (BT/PR30159/MED/15/188/2018) and BT/PR40384/COT/142/5/2020.

## Supplementary Materials

### Supplementary Material 1

Methodology for immunofluorescence assays for antibodies against human coronaviruses OC43, NL63 and 229E.

### Cell lines and virus

LLC MK2 cells were from European Collection of Authenticated Cell Cultures Salisbury, UK (ECACC - 85062804). Baby Hamster Kidney (BHK-21) cells were from American Type Culture Collection, Manassas, VA, USA (ATCC-CCL-10). Cells were grown at 37°C in Minimum Essential medium (MEM) (Gibco-11090-073) containing 10% heat-inactivated fetal bovine serum (FBS) (Gibco-16140-071), 100 U/mL of penicillin and 100 µg/mL of streptomycin (Gibco-10378-016). Human hepatoma cells (Huh-7) (Japanese Collection of Research Bioresources Cell Bank) were grown at 37° C in Dulbecco’s minimum essential medium (DMEM) (Lonza 12-707F) containing above additives and non-essential amino acids (Gibco-111400-50). Virus isolates were obtained through BEI Resources, NIAID, NIH: Human Coronavirus, 229E, NR-52726; Human Coronavirus, NL63, NR-470; Human Coronavirus, OC43, NR-52725.

### Virus propagation

HCoV-OC43 was propagated in BHK-21 and HCoV-NL63 in LLC MK2 cells in MEM supplemented with 2% heat-inactivated FBS, 100 U/mL of penicillin, and 100 µg/L of streptomycin at 34° C under a humidified atmosphere of 5% CO2 for 4 days and 6 days respectively. HCoV-229E was propagated in Huh-7 cells in DMEM supplemented with 2% heat-inactivated FBS and other additives at 34° C for 2 days. Cells were observed for cytopathic effect and viruses were harvested by scraping the cells and resuspending in media. The cell suspensions were subjected to two rounds of freeze-thaw cycles by dipping the tubes containing cell suspensions sequentially in liquid nitrogen and 37° C water bath. Cell lysates were centrifuged for 10 min at 4000 rpm. Cleared supernatant was aliquoted and stored at - 70°C. LLC MK2 and Huh-7 cells were seeded at a density of 30,000 and 50,000 cells/well respectively in wells of 48-well plates. LLC MK2 cells were infected with HCoV-NL63 and HCoV-OC43 and Huh-7 cells were infected with HCoV-229E. The cells were infected with serial 10-fold dilutions of virus stocks (10^−1^ to 10^−5^) prepared in MEM or DMEM supplemented with 2% heat-inactivated FBS, 100 U/mL of penicillin, and 100 µg/mL of streptomycin at 34°C under a humidified atmosphere of 5% CO_2_ for 48 hours. After 48 hours post-infection, the infected cells were quantified by immunofluorescence assay as described in the next section. Virus dilutions of 1:10 for HCoV-NL63 and HCoV-OC43 infection in LLC MK2 and 1:10000 for HCoV-229E infection in Huh-7 cells were determined as optimum for infection without cytopathic effect.

### Immunofluorescence

48 h post-infection cells were washed with cold PBS twice and fixed with cold methanol at - 20°C for 20 min. Cells were washed twice with PBS and three times with PBS containing 0.2% BSA (PBS-BSA) followed by incubation with primary (one hour) and secondary antibodies (30 min). The primary antibody used was either human sera from patients (diluted 1:10 in PBS containing 0.2% BSA) or a commercial rabbit polyclonal antibody (1:500 dilution in PBS-BSA) against the nucleocapsid (Catalog nos. 40640/41/43-T62) obtained from Sino Biologicals, Beijing, China which was used a positive control. Cells were washed three times with PBS-BSA after one hour incubation at room temperature. 1:500 dilution of secondary antibody in PBS-BSA was used. The secondary antibodies used were goat anti-human IgG-Alexa 488 (A11013 - ThermoFisher) for human sera or goat anti-rabbit IgG-Alexa 488 (A21206-ThermoFisher) for nucleocapsid antibodies. Cells were washed after respective antibody incubations with PBS-BSA three times. Nuclei were stained by Hoechst stain (1:10000 dilution in PBS) for 5 min and washed twice with PBS. Images were acquired using IX83 fluorescence microscope (Olympus).

## Supplementary Figures

**Supplementary Figure 1.**
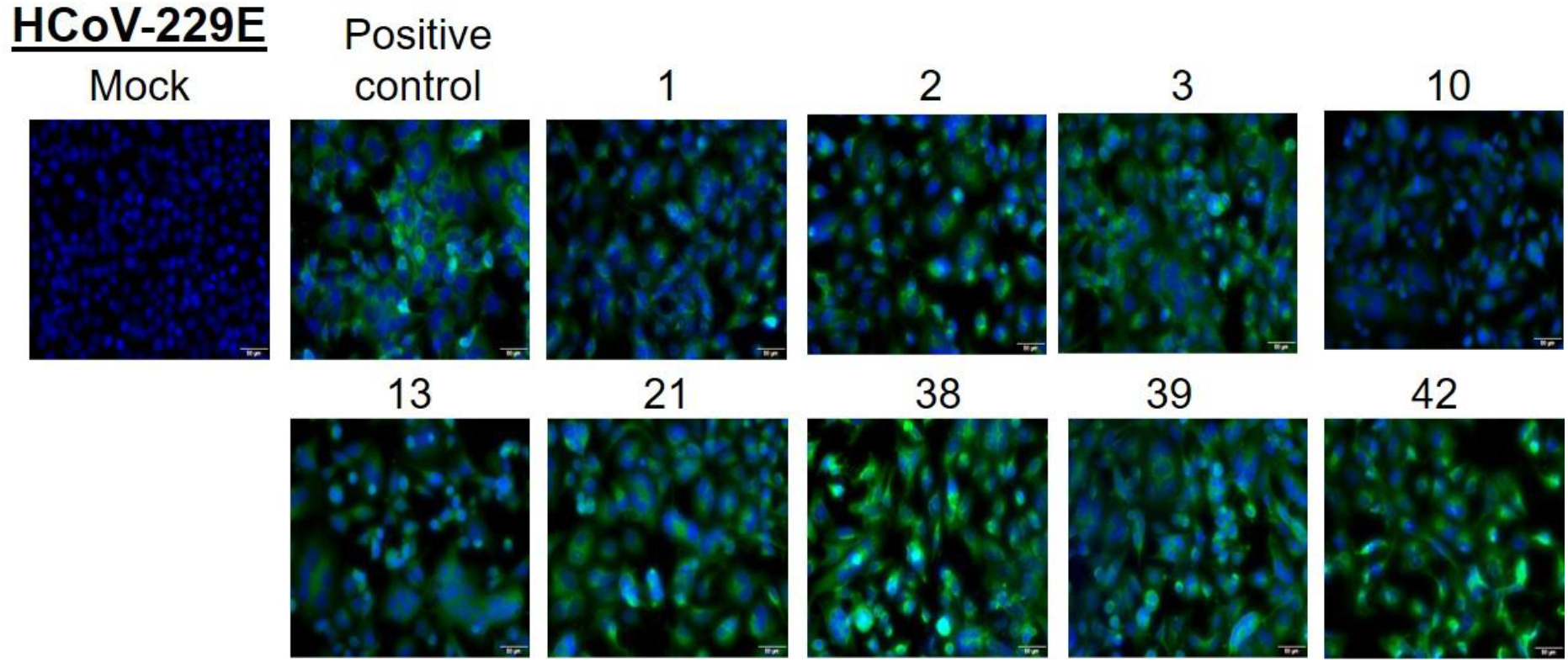
LLC MK2 cells were infected with HCoV-OC43 and at 48 h post-infection, cells were fixed and stained with 1:10 dilution of serum samples from indicated patients. A commercial antibody against the N protein is used as a positive control. Alexa-488 conjugated secondary antibody (either anti-human or anti-rabbit) was used to detect the primary antibody. Nuclei were stained by Hoechst stain. Scale 10 μm.

**Supplementary Figure 2.**
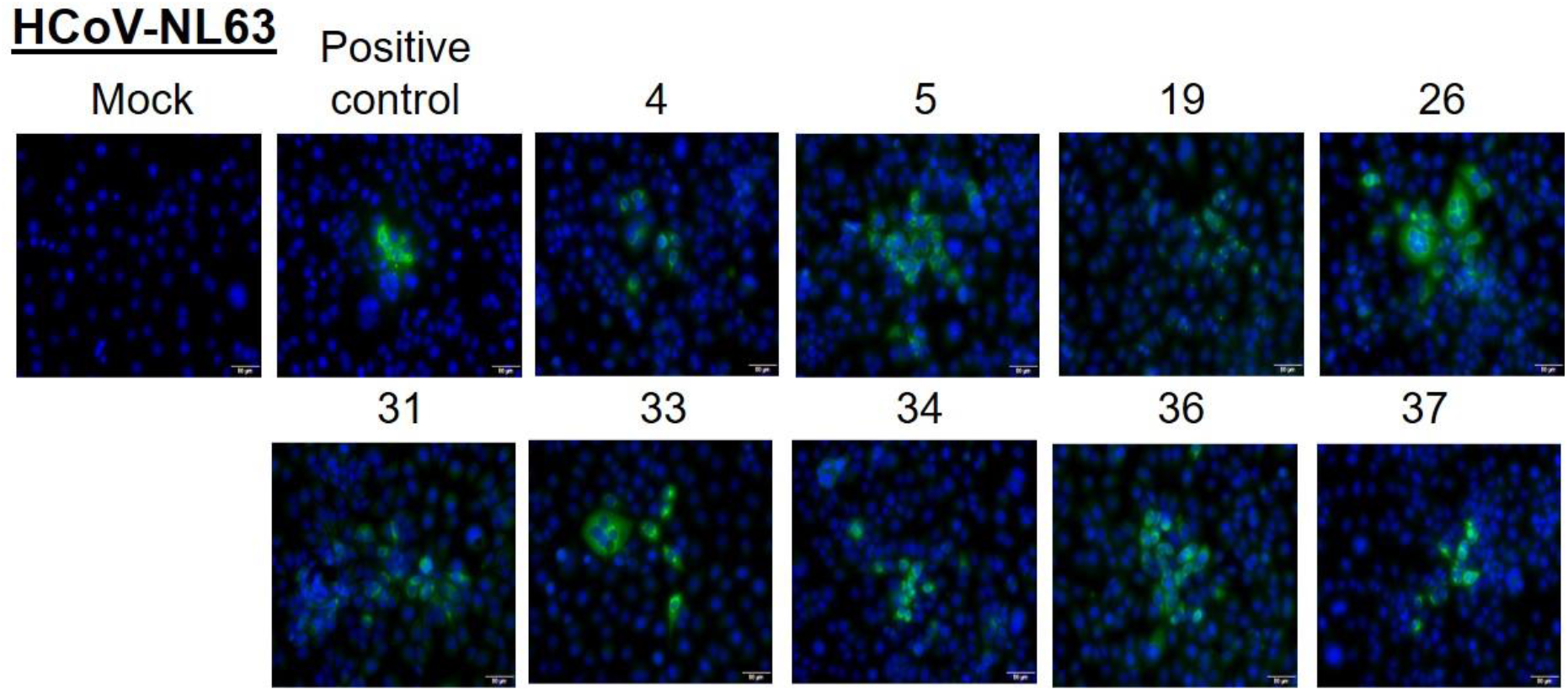
LLC MK2 cells were infected with HCoV-NL63 and at 48 h post-infection, cells were fixed and stained with 1:10 dilution of serum samples from indicated patients. A commercial antibody against the N protein is used as a positive control. Alexa-488 conjugated secondary antibody (either anti-human or anti-rabbit) was used to detect the primary antibody. Nuclei were stained by Hoechst stain. Scale 10 μm.

**Supplementary Figure 3.**
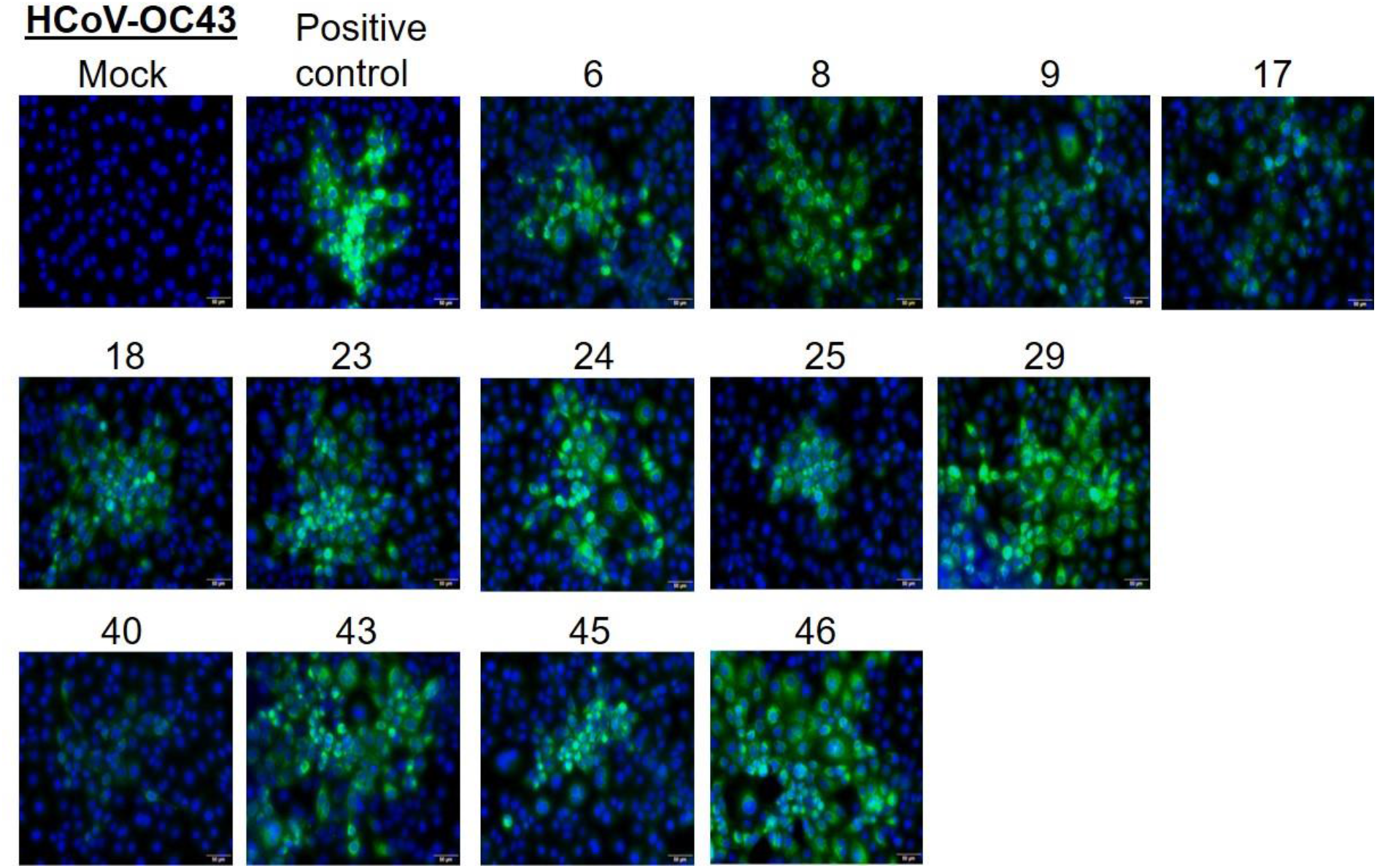
Huh-7 cells were infected with HCoV-229E and at 48 h post-infection, cells were fixed and stained with 1:10 dilution of serum samples from indicated patients. A commercial antibody against the N protein is used as a positive control. Alexa-488 conjugated secondary antibody (either anti-human or anti-rabbit) was used to detect the primary antibody. Nuclei were stained by Hoechst stain. Scale 10 μm.

**Supplementary Figure 4.**
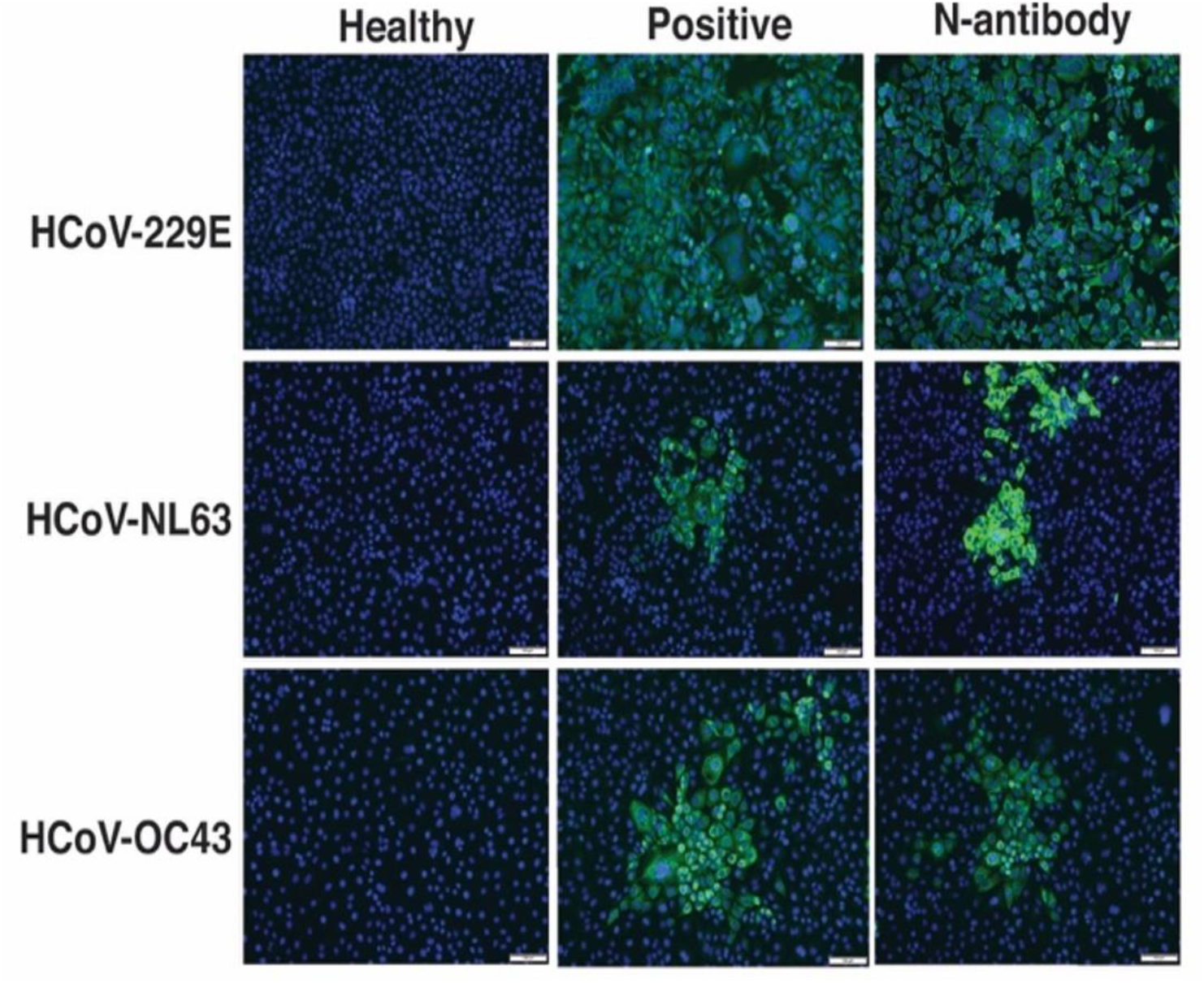
Huh-7 cells were infected with HCoV-229E and LLC MK2 cells were infected with HCoV-NL63 or HCoV-OC43 and at 48 h post-infection, cells were fixed and stained with 1:10 dilution of serum samples from a healthy or a positive subject. A commercial antibody against the N protein is used as an assay positive control. Alexa-488 conjugated secondary antibody (either anti-human or anti-rabbit) was used to detect the primary antibody. Nuclei were stained by Hoechst stain.

## References

1. Kumar P, Mukherjee A, Randev S, et al. Epidemiology of coronavirus infection in children and their impact on lung health: Finding from a birth cohort study. Pediatr Infect Dis J. 2020;39:e452–4.

2. Anderson EM, Goodwin EC, Verma A, et al. Seasonal human coronavirus antibodies are boosted upon SARS-CoV-2 infection but not associated with protection. Cell. 2021: 184(7):1858–1864.e10.

3. Poston D, Weisblum Y, Wise H, et al. Absence of SARS-CoV-2 neutralizing activity in pre-pandemic sera from individuals with recent seasonal coronavirus infection. Clin Infect Dis. 2020:ciaa1803. doi: 10.1093/cid/ciaa1803

4. Kumar P, Mukherjee A, Randev S, et al. Effect of acute respiratory infections in infancy on pulmonary function test at 3 years of age: a prospective birth cohort study. BMJ Open Respir Res. 2020;7:e000436.

5. Anantharaj A, Gujjar S, Kumar S, et al. Kinetics of viral load, immunological mediators and characterization of a SARS-CoV-2 isolate in mild COVID-19 patients during acute phase of infection. medRxiv, 2020. https://www.medrxiv.org/content/10.1101/2020.11.05.20226621v2. Accessed 18 February 2021.

6. Bewley KR, Coombes NS, Gagnon L, et al. Quantification of SARS-CoV-2 neutralizing antibody by wild-type plaque reduction neutralization, microneutralization and pseudotyped virus neutralization assays. Nat Protoc, 2021; 16: 3114–3140.

7. Mateus J, Grifoni A, Tarke A, et al. Selective and cross-reactive SARS-CoV-2 T cell epitopes in unexposed humans. Science. 2020;370:89–94.

